# Wealth inequalities malaria parasitaemia and child anaemia in Nigeria

**DOI:** 10.64898/2026.11.08.26367570

**Authors:** Kayode Raphael Fowobaje, Abiodun Adegbenro, Tombotamunoa Lawson

## Abstract

Anaemia and malaria remain concentrated among children living in poorer households in Nigeria, but the extent to which the wealth gradient in anaemia operates through current malaria parasitaemia is uncertain. We used three nationally representative surveys to estimate a clearly defined parasitaemia-mediated component of the association, while recognising that anaemia has multiple infectious, nutritional, genetic, and social causes.

We analysed de facto children aged 6-59 months in the 2010, 2015, and 2021 Nigeria Malaria Indicator Surveys. Within each survey, we contrasted children in the poorest quintile with children in the richer and richest quintiles. The outcome was altitude-adjusted anaemia and the mediator was microscopy-confirmed parasitaemia. Weighted outcome and mediator models adjusted for child age and sex, maternal education, urban residence, geopolitical zone, household size, rainy-season interview, and altitude. We standardized interventional total, direct, and indirect effects using g- computation. Missing data were addressed with 20 chained-equation imputations; uncertainty used 1,000 Rao-Wu survey-bootstrap replicates.

Analytic samples included 3,307, 3,643, and 6,572 children in 2010, 2015, and 2021. The multiple-imputation total risk difference was 11.0 (95% CI 4.1 to 17.9) percentage points in 2015 and 12.5 (95% CI 8.4 to 16.6) points in 2021; corresponding parasitaemia-mediated indirect differences were 3.4 (95% CI 1.4 to 5.4) and 2.0 (95% CI 1.1 to 2.9) points. The 2010 total-effect interval included zero. Proportion-mediated estimates were withheld for 2010 and 2015 because of unstable total- effect denominators.

The poorest-to-richer/richest contrast was associated with higher anaemia risk in 2015 and 2021. A smaller positive component was compatible with mediation through microscopy-confirmed parasitaemia. Because wealth, parasitaemia, and haemoglobin were measured in the same survey visit, the estimates require assumptions about temporal ordering and unmeasured mediator-outcome confounding; they do not establish that eliminating parasitaemia would remove the observed anaemia inequality.

## Introduction

Malaria and anaemia commonly co-occur among young children in Nigeria, with substantial geographic and socioeconomic variation. Earlier Nigerian work has modelled their joint spatial distribution, but it did not estimate an exposure-specific pathway from household wealth through laboratory-confirmed parasitaemia to anaemia [1]. A recent systematic review found that formal mediation analyses of socioeconomic disparities in malaria are uncommon and often limited by cross- sectional measurement, incomplete confounding control, and inadequate handling of exposure- mediator interaction [2].

Household wealth may influence malaria risk through housing, prevention, access to care, and other pathways. Malaria parasitaemia can in turn contribute to anaemia, but nutritional deficiencies, helminths, inherited haemoglobin disorders, and other causes mean that malaria cannot account for all anaemia. The study objective was to quantify standardized interventional total, direct, and parasitaemia-mediated effects of the poorest household wealth category on child anaemia, separately in each NMIS round.

Recent Nigerian analyses continue to show marked heterogeneity in childhood malaria risk by place, household circumstances, prevention coverage, and season [3]. National trend work found that reductions in infection were not uniform across states and zones, and recent analyses of the 2021 survey have likewise identified geographically patterned risk [4,5,6]. Studies of care seeking, antimalarial use, malaria knowledge, seasonal chemoprevention, malaria communication, vaccine readiness, parasite clearance, and programme delivery also illustrate that the social conditions surrounding exposure and response do not reduce to a single household asset score [7–19]. The present study therefore does not treat wealth as a manipulable intervention. It treats wealth as a summary exposure whose association with anaemia may operate through several pathways.

A mediation analysis adds information only if the causal question and the resulting assumptions are explicit. We chose microscopy-confirmed parasitaemia as a biologically proximal mediator, rather than using reported fever or treatment. That choice is supported by contemporary malaria research in Nigeria and elsewhere, but it also narrows the interpretation: a negative microscopy result does not exclude recent infection, and a positive test is not a complete measure of cumulative malaria burden [5,6,20–24]. We estimate an interventional indirect effect through the measured parasite status, not the proportion of anaemia biologically caused by malaria.

The primary objective was to estimate, within each survey round, the difference in anaemia risk had the observed study population experienced the mediator distribution expected under the poorest versus the comparison wealth categories. Secondary objectives were to compare the resulting estimates with complete-case analyses and to describe temporal changes in the total, direct, and indirect components. We report the work as an observational, design-based analysis in accordance with STROBE and current guidance for reporting mediation analyses [25,26].

The study is deliberately framed as a repeated cross-sectional analysis rather than as a trend analysis that pools survey years. The relationship between household wealth, malaria exposure, prevention, nutritional conditions, and anaemia can vary with transmission intensity and survey implementation. Estimating effects separately by year avoids assuming that an identically labelled wealth quintile or covariate has the same meaning or distribution in 2010, 2015, and 2021. It also avoids treating secular change as an individual longitudinal trajectory. Comparisons between years are therefore descriptive comparisons of standardized quantities, not estimates of within-child change.

We distinguish the central epidemiologic question from a policy simulation. The exposure contrast compares observed social positions; it does not describe an intervention that can simply move a household from the poorest category to a richer category while holding all other social circumstances fixed. The interventional mediator distribution is consequently used as a formal device to separate a component compatible with current parasitaemia from a component that remains outside that measured pathway. This makes the estimand more transparent, but it does not remove the requirement for consistency, positivity, correct model specification, and no unmeasured confounding of the relevant relationships.

## Methods

### Design and data source

We used the nationally representative Nigeria Malaria Indicator Surveys conducted in 2010, 2015, and 2021 [27]. The analytic population comprised de facto household members aged 6-59 months. The pipeline retained DHS sampling weights, primary sampling units, and strata. Children outside the age range and records without a valid outcome in complete-case analyses were excluded. The participant flow is reported in Supplementary Table S1. The public-use survey datasets were accessed for research purposes on September 15, 2026. The authors did not have access to information that could identify individual participants during or after data collection.

### Variables

The exposure was household wealth, defined within survey as the poorest quintile versus the richer and richest quintiles combined; middle categories were excluded from this prespecified contrast. The outcome was any altitude-adjusted anaemia, defined from HC57 categories 1-3. The mediator was microscopy-confirmed malaria parasitaemia (HML32=1). Covariates were child age modelled using a natural cubic spline, sex, maternal education, urban residence, six geopolitical zones, household size, rainy-season interview, and altitude modelled using a natural cubic spline. These variables were selected as measured pre-exposure or contextual factors. ITN use was treated as a post-wealth pathway variable rather than a confounder in the single-mediator estimand.

### Causal structure and confounding control

The prespecified directed acyclic graph is supplied as Figure S1 in the supplementary appendix. It represents geographic context, season, altitude, maternal education, child age and sex, and household composition as common causes or pre-exposure contextual determinants of wealth, parasitaemia, and anaemia. These variables were included in both mediator and outcome models. Household net ownership, ITN use, housing quality, and treatment access were not included as mediator-outcome confounders because they can plausibly lie downstream of wealth and would change the target estimand if conditioned upon. The DAG is a transparent statement of assumptions rather than a claim that all confounding was measured [28].

Anaemia was defined as any altitude-adjusted haemoglobin category below the no-anaemia category in the survey recode. We did not attempt to reclassify anaemia by presumed aetiology. Maternal anaemia, child diet, helminth infection, haemoglobinopathy, recent antimalarial treatment, and individual mosquito exposure were either unavailable or not harmonised across all rounds. They are consequently potential sources of residual confounding or competing mechanisms, and their absence is reflected in the causal interpretation rather than obscured by a data-driven variable-selection procedure.

### Statistical analysis

For each survey, weighted logistic models were fitted for parasitaemia and anaemia; the latter included wealth, parasitaemia, their interaction, and the prespecified covariates. Analytic g- computation standardized predictions over the observed analytic population. We report interventional total effects, interventional direct effects, and interventional indirect effects through parasitaemia as risk differences and risk ratios. This estimand accommodates exposure-mediator interaction and avoids interpreting a conventional regression product as a causal indirect effect [25,29,30].

The primary analysis used 20 survey-specific chained-equation imputations. Microscopy, RDT, altitude-adjusted haemoglobin, ITN use, and household net ownership were included in the imputation procedure, with anaemia passively derived from haemoglobin. Supplementary Tables S2 and S3 document variable missingness and imputation diagnostics. Within-survey uncertainty used 1,000 PSU-within-stratum survey-bootstrap replicates distributed across imputations, with within- and between-imputation uncertainty combined using Rubin’s rules [31–33]. Confidence intervals are 95% intervals; mediation contrasts were not assigned null-hypothesis p-values.

Models were fitted separately by survey because wealth quintiles, field conditions, and the joint distributions of determinants differed across time. Sampling weights were scaled to the analytic sample and survey strata and primary sampling units were retained in every bootstrap replicate. The resampling procedure drew PSUs within strata and recalculated all fitted models and standardized quantities; it did not merely resample final estimates. This approach reflects both imputation and sampling uncertainty, although the resulting intervals do not capture bias from an incorrect causal graph or unmeasured confounding [31–33].

We assessed positivity descriptively by cross-tabulating exposure, mediator, and key covariates within survey and did not extrapolate standardized predictions outside observed covariate support. The data-cleaning and analysis scripts specify the survey recodes, derived variables, model formulae, imputation predictor matrices, and bootstrap seed. Analyses used R and the survey, mice, and mediation-supporting packages; exact software versions are retained in the reproducibility record. No sample-size calculation was undertaken because the analysis used all eligible children in each public-use survey.

### Effect measures and interpretation

The standardized total effect contrasts predicted anaemia risks under poorest versus comparison household wealth. The interventional direct and indirect effects partition that contrast by a stochastic intervention on the microscopy-confirmed mediator distribution, conditional on the specified models. On the risk-difference scale, total = direct + indirect to numerical precision. Proportions mediated are reported only when total-effect denominator stability is supported by bootstrap checks. Thus the 2010 ratio (total-effect CI includes zero) and 2015 ratio (one bootstrap total-effect sign reversal) were withheld, including their confidence intervals; the absolute effects remain reportable.

For each imputed data set, continuous age and altitude were entered with the same prespecified spline specification in the mediator and outcome models. Categorical predictors retained a common reference coding within a survey, and the wealth-by-mediator interaction was retained regardless of its estimated magnitude because it is needed for the stated interventional interpretation. Model predictions were averaged over the observed covariate distribution rather than evaluated at a hypothetical average child. This standardization makes the absolute risks interpretable for the analytic survey population and prevents covariate patterns from being represented by an artificial reference profile.

We inspected convergence plots and marginal distributions for the imputation models and checked that imputed binary values remained within the permissible recode range. The supplementary appendix documents the variables entered into imputation, the passive derivation of anaemia, and comparison of complete-case and imputed estimates. Missingness was not assumed to be completely at random; the chained-equation approach instead relies on a missing-at-random assumption conditional on included predictors. It cannot correct for systematic absence related to unmeasured haemoglobin, parasite burden, treatment, or social conditions.

## Results

The multiple-imputation samples comprised 3,307, 3,643, and 6,572 children in 2010, 2015, and 2021, respectively (Supplementary Table S1). In 2021, weighted anaemia prevalence was 79.9% among the poorest children and 54.0% among the richest; microscopy positivity was 31.3% and 5.3%, respectively (Table 1).

**Table 1.**
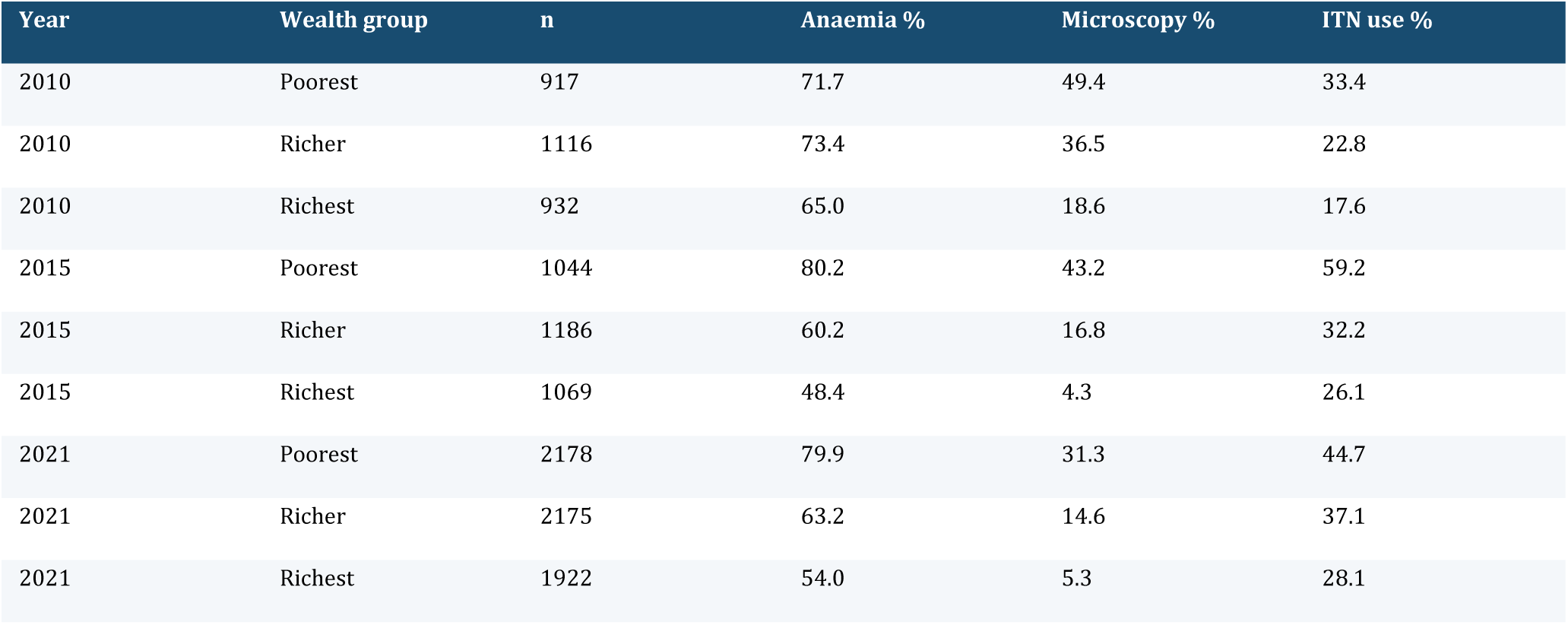
Weighted descriptive prevalence by survey-specific household wealth quintile.

In 2010, the standardized total risk difference was 1.5 (95% CI -6.3 to 9.3) percentage points; the indirect difference through microscopy-confirmed parasitaemia was 1.9 (95% CI 0.5 to 3.4) points. Because the total-effect confidence interval crossed zero, the proportion mediated and its confidence interval were withheld (Table 2; Figure 1).

**Figure 1.**
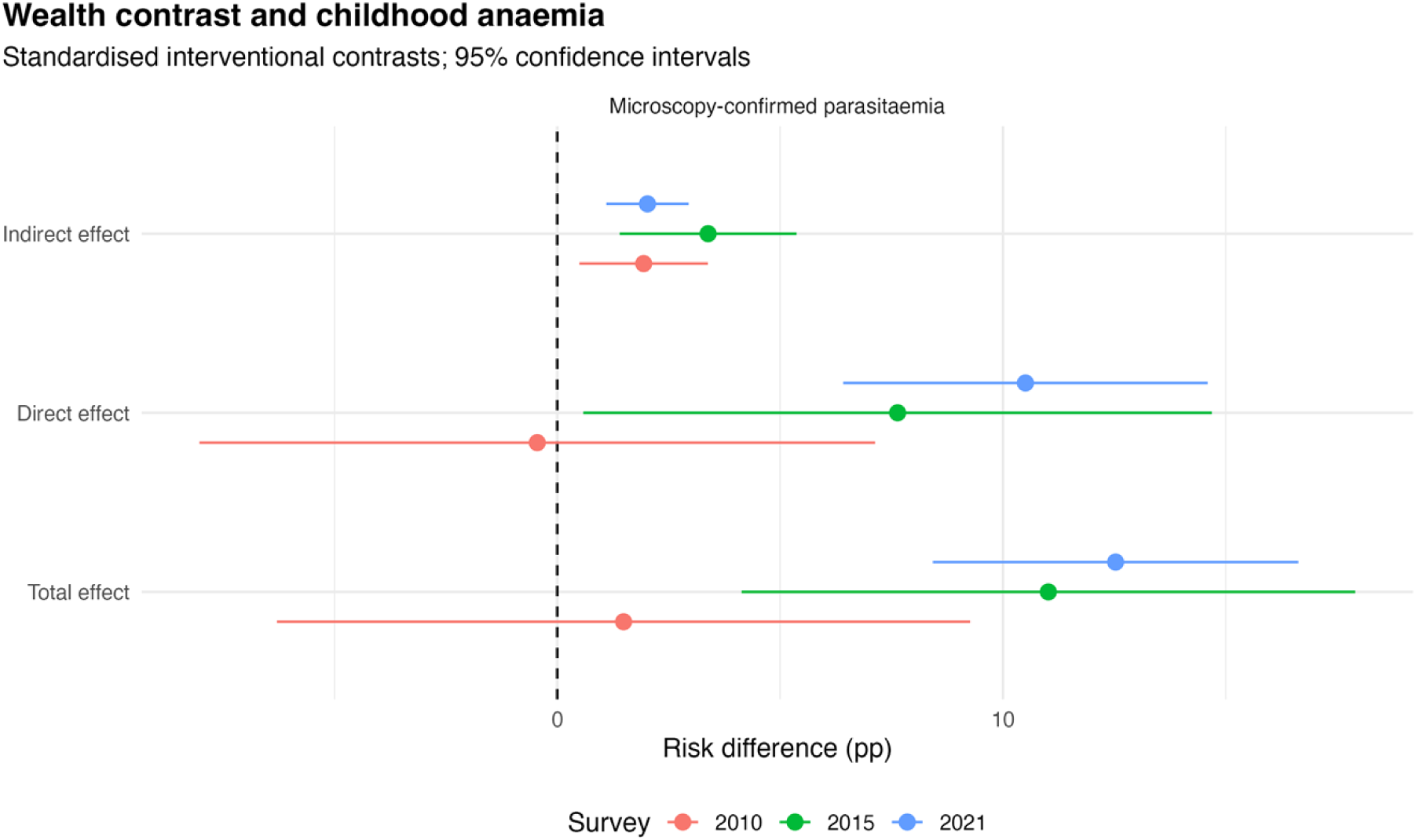
Multiple-imputation standardized total, direct, and parasitaemia-mediated risk differences by survey year with 95% confidence intervals.

**Table 2.** Multiple-imputation interventional contrasts for poorest versus richer/richest household wealth through microscopy-confirmed parasitaemia. Risk differences are percentage points and risk ratios are unitless. A dash denotes proportion mediated deliberately withheld for denominator instability (2010 and 2015).

| **Year** | **Quantity** | **Estimate** | **95% CI** |
|---|---|---|---|
| 2010 | TE_rd | 1.49 | -6.29 to 9.26 |
| 2010 | IDE_rd | -0.45 | -8.03 to 7.13 |
| 2010 | IIE_rd | 1.94 | 0.50 to 3.38 |
| 2010 | PM | Withheld | — |
| 2010 | TE_rr | 1.02 | 0.91 to 1.13 |
| 2010 | IIE_rr | 1.03 | 1.01 to 1.05 |
| 2015 | TE_rd | 11.02 | 4.13 to 17.90 |
| 2015 | IDE_rd | 7.64 | 0.58 to 14.69 |
| 2015 | IIE_rd | 3.38 | 1.40 to 5.37 |
| 2015 | PM | Withheld | — |
| 2015 | TE_rr | 1.18 | 1.06 to 1.30 |
| 2015 | IIE_rr | 1.05 | 1.02 to 1.08 |
| 2021 | TE_rd | 12.53 | 8.43 to 16.63 |
| 2021 | IDE_rd | 10.50 | 6.41 to 14.59 |
| 2021 | IIE_rd | 2.02 | 1.10 to 2.95 |
| 2021 | PM | 16.17 | 7.00 to 25.34 |
| 2021 | TE_rr | 1.20 | 1.13 to 1.27 |
| 2021 | IIE_rr | 1.03 | 1.01 to 1.04 |

In 2015, the total risk difference was 11.0 (95% CI 4.1 to 17.9) percentage points, comprising a direct component of 7.6 (95% CI 0.6 to 14.7) and a parasitaemia-mediated indirect component of 3.4 (95% CI 1.4 to 5.4) points. Total and indirect risk ratios were 1.18 and 1.05. The 2015 proportion-mediated point estimate and confidence interval are deliberately not reported: a near-zero total-effect denominator and a bootstrap sign reversal made the ratio unstable (one of 1,000 replicates; Table 2).

In 2021, the total risk difference was 12.5 (95% CI 8.4 to 16.6) percentage points, comprising direct and indirect components of 10.5 (95% CI 6.4 to 14.6) and 2.0 (95% CI 1.1 to 2.9) points. The proportion mediated was 16.2 (95% CI 7.0 to 25.3)% and met the stability checks (Table 2; Figure 2).

**Figure 2.**
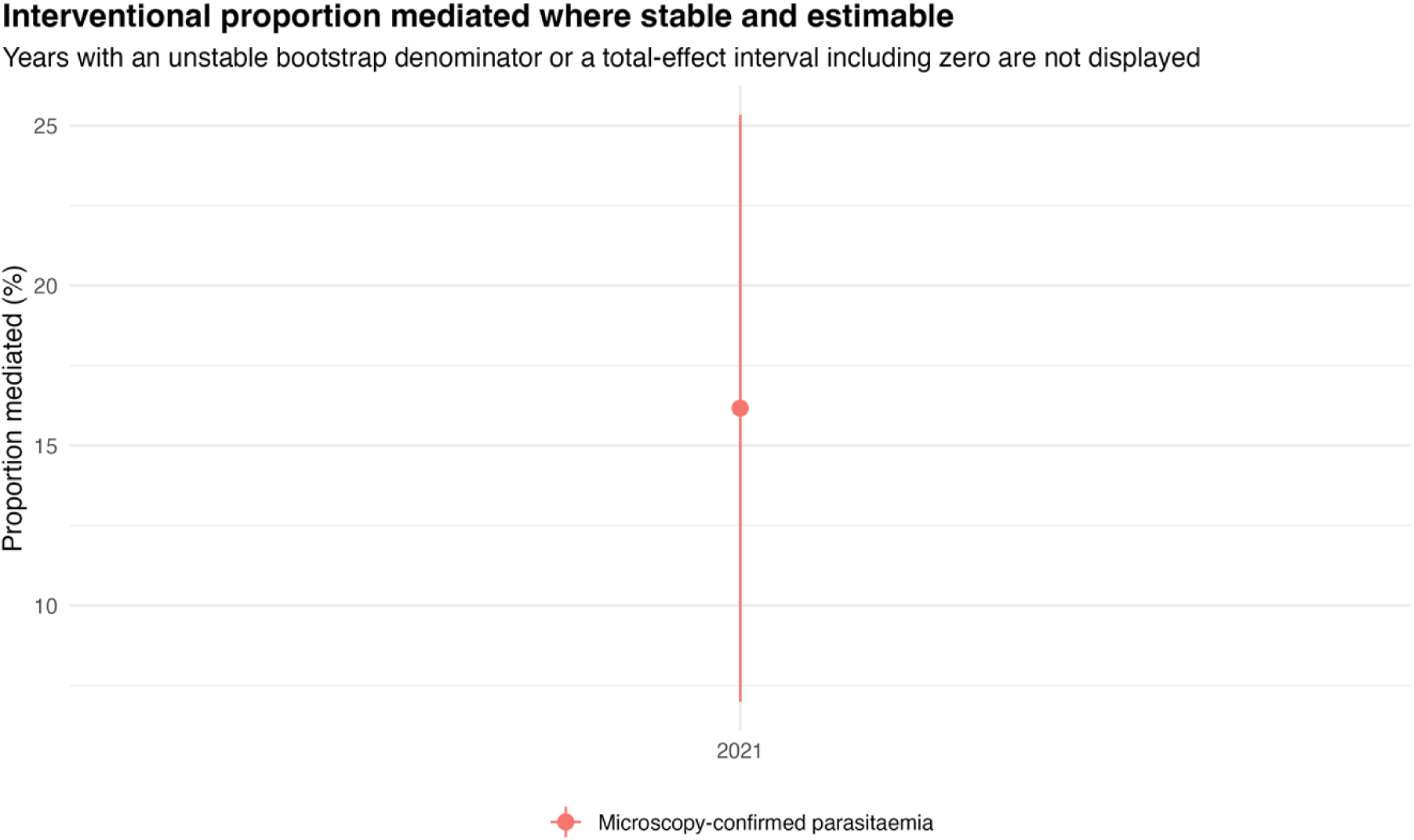
Proportion mediated in 2021 only (multiple imputation; 95% confidence interval). The 2010 and 2015 ratio estimates and intervals failed the denominator-stability criteria and are not displayed.

Complete-case and multiple-imputation estimates were compared for the total, direct, and indirect risk differences (Supplementary Table S5; Figure S2). In 2015, total differences were 10.8 and 11.0 percentage points, respectively, with indirect differences of 3.1 and 3.4 points. In 2021, total differences were approximately 12.4 and 12.5 points, and indirect differences were approximately 2.0 points using either method. The 2010 total difference was imprecise with both methods.

The parasitaemia-associated indirect component was positive in all three rounds, but the overall wealth-associated anaemia contrast was estimated imprecisely in 2010. In 2015 and 2021, total and indirect risk ratios were above one (Table 2); absolute contrasts are emphasized to retain a public-health scale. Bootstrap intervals describe uncertainty under the stated survey and imputation procedures and do not by themselves validate a causal pathway.

The descriptive gradients differed by survey. In 2010, microscopy positivity was 49.4% in the poorest group and 18.6% in the richest group, but anaemia was common across wealth categories. The 2015 microscopy gradient was 43.2% versus 4.3%, and the 2021 gradient was 31.3% versus 5.3%. These estimates are weighted descriptive prevalences rather than standardized causal effects (Table 1; Supplementary Table S6).

In 2015, the absolute parasitaemia indirect contrast (3.4 percentage points) was smaller than the direct contrast (7.6 points); the corresponding 2021 contrasts were 2.0 and 10.5 points. Much of the standardized wealth-associated anaemia difference lay outside the measured parasite pathway.

Secondary joint ITN–parasitaemia contrasts are reported in Supplementary Table S8; they are not estimates of an isolated sequential ITN-to-parasitaemia mechanism.

The indirect risk-difference confidence intervals excluded zero in 2015 and 2021 under the fitted models and bootstrap procedures. Only the 2021 proportion-mediated estimate passed denominator- stability checks; the 2010 and 2015 proportions and intervals are omitted from the manuscript, tables, and figures. Missingness, imputation, and survey-recode diagnostics appear in Supplementary Tables S2, S3, and S7 and Figure S3.

## Discussion

The wealth contrast was associated with higher anaemia risk in 2015 and 2021. The indirect component through microscopy-confirmed parasitaemia was smaller than the total contrast but consistently positive. These estimates align with evidence that socioeconomic position is associated with malaria in sub-Saharan Africa and that malaria-related pathways are plausible, while also recognising that direct evidence on mediation remains limited [2].

The large direct component is consistent with the multifactorial causes of childhood anaemia. The analysis was not designed to attribute all anaemia to malaria. Nutrition, helminths, inherited haemoglobin conditions, maternal anaemia, recent treatment, and individual environmental exposure were not comparably measured as pre-mediator confounders in all rounds. The findings should therefore be interpreted as standardized, assumption-dependent mediation estimates rather than evidence that changing wealth or parasitaemia would necessarily change anaemia at the same magnitude.

The results complement, but do not duplicate, descriptive studies of Nigerian malaria trends. Prior work has documented reductions in infection and continuing geographic heterogeneity [4–6]. Other recent studies link household and caregiver conditions to infection prevention, care seeking, and treatment [7–12]. Our contribution is to distinguish a measured parasite pathway from the residual wealth-associated difference in anaemia. The modest indirect component is compatible with a role for malaria control in reducing inequity, but it should not be converted into a statement that a fixed share of anaemia is preventable by malaria treatment or vector control.

The 2015 and 2021 findings are also consistent with the broader literature on poverty and malaria, which identifies multiple pathways connecting material circumstances to exposure, prevention, diagnosis, treatment, and recovery [2,20]. In that framework, a small-to-moderate parasitaemia- mediated component is plausible because a cross-sectional test measures one point in a dynamic infection history and because many determinants of anaemia do not act through detectable parasitaemia. Recent Nigerian research on spatial prediction and clinical febrile illness similarly cautions against representing malaria risk as homogeneous within a national survey [5,6,23,24].

The main causal limitations are substantial. Cross-sectional data cannot establish that the wealth contrast preceded the mediator, that the mediator preceded haemoglobin status, or that measured covariates control all exposure-outcome, exposure-mediator, and mediator-outcome confounding. Exposure-induced mediator-outcome confounders would also violate standard identification conditions. Laboratory classification, seasonal timing, and selection into biomarker testing may introduce additional error. The confidence intervals quantify sampling and imputation uncertainty conditional on the specified models; they are not uncertainty intervals for these causal assumptions.

The study has several strengths: use of microscopy rather than self-reported malaria as the mediator; prespecified, survey-harmonised confounder adjustment; explicit handling of exposure-mediator interaction; multiple imputation; and 1,000 survey-respecting bootstrap replicates. Future longitudinal studies should collect repeated parasite and haemoglobin measures, diet, inherited haemoglobin conditions, treatment histories, household environmental measures, and fine-scale vector exposure. Such data would allow stronger temporal ordering and a more complete multiple-mediator analysis.

Until then, the present estimates are best used to frame hypotheses and to describe the potential importance of the parasite pathway alongside wider determinants of childhood anaemia.

The temporal contrast between 2015 and 2021 should be interpreted carefully. A larger absolute total effect in 2021 does not demonstrate a widening causal effect of wealth, because the analysis uses round-specific wealth categories and the underlying prevalence, diagnostic conditions, household composition, and contextual covariates differ across rounds. Conversely, a similar relative effect does not establish invariance of the pathway. Reporting each survey separately is intended to make these limits visible. A pooled model with a year interaction could be informative in a separate study, but would require stronger assumptions about harmonization and the comparability of the exposure measure.

We also did not interpret ITN use, household net ownership, housing quality, or treatment access as confounders of the wealth-parasitaemia relationship. They may be targets of policy and important determinants of malaria, but conditioning on variables downstream of wealth would estimate a different direct pathway and might introduce collider bias. The DAG makes that decision reviewable. Alternative diagrams that position one or more of these variables as pre-exposure determinants would imply a different estimand, which is why causal conclusions must remain conditional on the stated substantive assumptions rather than on the statistical procedure alone.

Finally, the analysis is relevant to equity monitoring because it makes clear that a reduction in parasitaemia need not remove the full anaemia disparity. Programmes addressing vector exposure, prompt diagnosis, and treatment may reduce the malaria-linked component. Parallel action on diet, maternal health, sanitation, helminth control, housing, and access to care is likely needed to address the residual difference. This is a conceptual implication of the multifactorial outcome, not a programme-effect estimate from these surveys. The result should be read alongside locally specific burden and implementation data when interventions are planned.

Strengths include microscopy-based parasitaemia, harmonized survey recodes, prespecified covariates, and combined imputation with survey-bootstrap inference. Limitations include simultaneous exposure, mediator, and outcome measurement; unmeasured mediator–outcome confounding; and susceptibility of ratio-based mediation summaries to small total effects. The 2015 proportion-mediated quantity is therefore suppressed rather than selectively interpreted. The joint- mediator contrast should not be assigned to a unique causal ITN-to-malaria sequence without further assumptions.

## Declarations

### Data Availability Statement

The data that support the findings of this study are available from the Demographic and Health Surveys (DHS) Program. The 2010, 2015, and 2021 Nigeria Malaria Indicator Survey (NMIS) datasets can be accessed by registered researchers upon request through the DHS Program website (https://dhsprogram.com/data/available-datasets.cfm). The authors confirm they had no special access privileges to these datasets that others would not have.

## Acknowledgements

The authors thank the Demographic and Health Surveys (DHS) Program, the National Population Commission, and ICF for granting access to the Nigeria Malaria Indicator Survey datasets used in this analysis.

## Ethics approval and consent to participate

This study is a secondary analysis of fully anonymized, public-use data from the 2015 and 2021 Nigeria Malaria Indicator Surveys (NMIS). The original survey protocols were reviewed and approved by the National Health Research Ethics Committee of Nigeria (NHREC) and the ICF Institutional Review Board. Informed consent was obtained from the parents or guardians of all participating children during the primary data collection. Because the present analysis relies exclusively on de- identified, secondary data, additional ethics approval and consent to participate are not applicable.

## Consent for publication

Not applicable.

